# A saliva-based RNA extraction-free workflow integrated with Cas13a for SARS-CoV-2 detection

**DOI:** 10.1101/2020.11.07.20227082

**Authors:** Iqbal Azmi, Md Imam Faizan, Rohit Kumar, Siddharth Raj Yadav, Nisha Chaudhary, Deepak Kumar Singh, Ruchika Butola, Aryan Ganotra, Gopal Datt Joshi, Gagan Deep Jhingan, Jawed Iqbal, Mohan C Joshi, Tanveer Ahmad

## Abstract

A major bottleneck in scaling-up COVID-19 testing is the need for sophisticated instruments and well-trained healthcare professionals, which are already overwhelmed due to the pandemic. Moreover, the high-sensitive SARS-CoV-2 diagnostics are contingent on an RNA extraction step, which, in turn, is restricted by constraints in the supply chain. Here, we present CASSPIT (**C**as13 **A**ssisted **S**aliva-based & Smartphone **I**ntegrated **T**esting), which will allow direct use of saliva samples without the need for an extra RNA extraction step for SARS-CoV-2 detection. CASSPIT utilizes CRISPR-Cas13a based SARS-CoV-2 RNA detection, and lateral-flow assay (LFA) readout of the test results. The sample preparation workflow includes an optimized chemical treatment and heat inactivation method, which, when applied to COVID-19 clinical samples, showed a 97% positive agreement with the RNA extraction method. With CASSPIT, LFA based visual limit of detection (LoD) for a given SARS-CoV-2 RNA spiked into the saliva samples was ∼200 copies; image analysis-based quantification further improved the analytical sensitivity to ∼100 copies. Upon validation of clinical sensitivity on RNA extraction-free saliva samples (n=76), a 98% agreement between the lateral-flow readout and RT-qPCR data was found (Ct<35). To enable user-friendly test results with provision for data storage and online consultation, we subsequently integrated lateral-flow strips with a smartphone application. We believe CASSPIT will eliminate our reliance on RT-qPCR by providing comparable sensitivity and will be a step toward establishing nucleic acid-based point-of-care (POC) testing for COVID-19.

## Introduction

Multi-step RNA extraction is a bottleneck that impedes mass testing for COVID-19. In this direction, RNA extraction-free assays are more suitable, which also provide a practical solution to develop point-of-care (POC) devices for genetic testing (Kriegova et al., 2020; Wee et al., 2020). Recently, RNA extraction-free methods were optimized on swab samples to detect SARS-CoV-2. The results show comparable sensitivity to RNA extraction methods (Alcoba-Florez et al., 2020; Brown et al., 2020; Bruce et al., 2020; Grant et al., 2020; Hasan et al., 2020; Merindol et al., 2020; Srivatsan et al., 2020; Wee et al., 2020). Similarly, these methods were also optimized on saliva samples (Lalli et al., 2020; Ranoa et al., 2020; Vogels et al., 2020b), although contradictory reports exist regarding sensitivity based on saliva than swab samples for SARS-CoV-2 (To et al., 2020; Williams et al., 2020; Wyllie et al., 2020; Meyerson et al., 2020). Nevertheless, all these studies unanimously suggest that nucleic acid extraction-free detection of SARS-CoV-2 is feasible.

These simple assay workflows have a tremendous potential to minimize the need for laboratory set-up and trained professionals, when integrated with a similar simplified method for detection. At present, the most robust and reliable detection method is based upon RT-qPCR, which is also a gold standard for COVID-19 testing. However, PCR-based detection methods have supply chain constraints to test on a large scale, and if available, may face a shortage of well-trained professionals to conduct the assay. Though, rapid POC tests which have been developed recently can perform mass testing of SARS-CoV-2 (Döhla et al., 2020; Jung et al., 2020) but, most of these tests are based on antigen/antibody detection and thus lack the sensitivity and specificity compared to genetic testing (Döhla et al., 2020). Recent advances in isothermal amplification-based assays provide a unique opportunity to detect nucleic acids under minimal instrument settings. These approaches, like RT-LAMP or RT-RPA, were developed previously and validated recently in SARS-CoV-2 containing clinical samples (Lalli et al., 2020; Piepenburg et al., 2006; Thai et al., 2004; Xia and Chen, 2020). Likewise, these isothermal-based amplification methods also have trade-offs in non-specific amplification (Zou et al., 2020). To circumvent these limitations, more robust methods based on CRISPR-Cas technology are employed, which utilizes collateral activities of Cas12 and Cas13 enzymes (Knott and Doudna, 2018; Li et al., 2019). These methods have been successfully used to detect human pathogens in various clinical samples, such as blood, saliva, and urine (Chen et al., 2018; Gootenberg et al., 2017). Cas12a works on DNA as input sample in a technique named DNA Endonuclease-Targeted CRISPR Trans Reporter (DETECTR). Recently, this technique was optimized to detect COVID-19 in swab samples with accuracy comparable to RT-qPCR (Broughton et al., 2020). Another such technique that detects single-stranded RNA is based on Cas13a, which is validated in many biological samples including saliva, and can reliably detect bacterial and viral pathogens with both LFA and fluorescent-based readout (Gootenberg et al., 2018; Gootenberg et al., 2017; Myhrvold et al., 2018). The technique called SHERLOCK (specific high-sensitivity enzymatic reporter unlocking) has a single-base specificity and single-molecule sensitivity with precision for multiplexing in a single reaction (Gootenberg et al., 2017). The SHERLOCK based diagnostics take advantage of extensive instrument free RPA or RT-RPA based pre-amplification of the nucleic acids, which makes this approach amenable and straightforward for on-site and home testing while at the same time providing better sensitivity and specificity (Gootenberg et al., 2018; Gootenberg et al., 2017; Myhrvold et al., 2018). Recently, SHERLOCK based diagnostics has been standardized and validated for the detection of SARS-CoV-2 in clinical swab samples (Patchsung et al., 2020).

Similarly, tools like All-in-One Dual CRISPR-Cas12a (AIOD-CRISPR) or colorimetric LAMP assay using Cas12a were developed (Ding et al., 2020; Joung et al., 2020). However, optimizing these methods utilized input RNA samples obtained using commercially available kits, which adds to the test’s cost and testing time. As of now, we have not come across any study which has used SHERLOCK based detection on RNA extraction-free clinical saliva samples for COVID-19 testing. In this study, we have clinically validated Cas13a integrated lateral-flow readout to detect SARS-CoV-2 in RNA extraction-free saliva samples. Further, we have developed a semi-quantitative method to provide high-sensitive test results of the lateral-flow test strip and integrated the test strip results with a smartphone application for field-deplorability and home testing.

## Results

### Optimization and validation of SARS-CoV-2 detection in clinical saliva samples

We used plasmids containing S and N genes of SARS-CoV-2, respectively, to standardize RT-qPCR, using CDC-approved and in-house designed primers. Among a set of 8 primers tested, two primer pairs for S gene (S-P-1, S-P-4) and CDC verified primer for N gene (N1) generated a single amplicon, with S gene amplification slightly better than N1 at same plasmid DNA concentration. RT-qPCR further confirmed these results (Figure 1A; Figure S1; Table S1). To determine the limit of detection (LoD), we generated S gene synthetic fragments containing T7 polymerase corresponding to the region flanking amplified sequence by S-P-1 primer. The synthetic DNA fragments were subsequently converted to RNA using an in-vitro transcription assay, following which the transcribed RNA was extracted, purified, and quantified (see Methods). We performed an RT-qPCR reaction with various purified RNA dilutions and plotted the corresponding Ct values against the known concentration (Figure 1B). Using S-P-1 primer and probe pair, we detected up to a single copy of RNA of S gene corresponding to Ct value <39.26. This LoD obtained for S gene agrees with the analytical sensitivity defined for the N gene (Vogels et al., 2020a).

**Figure 1:**
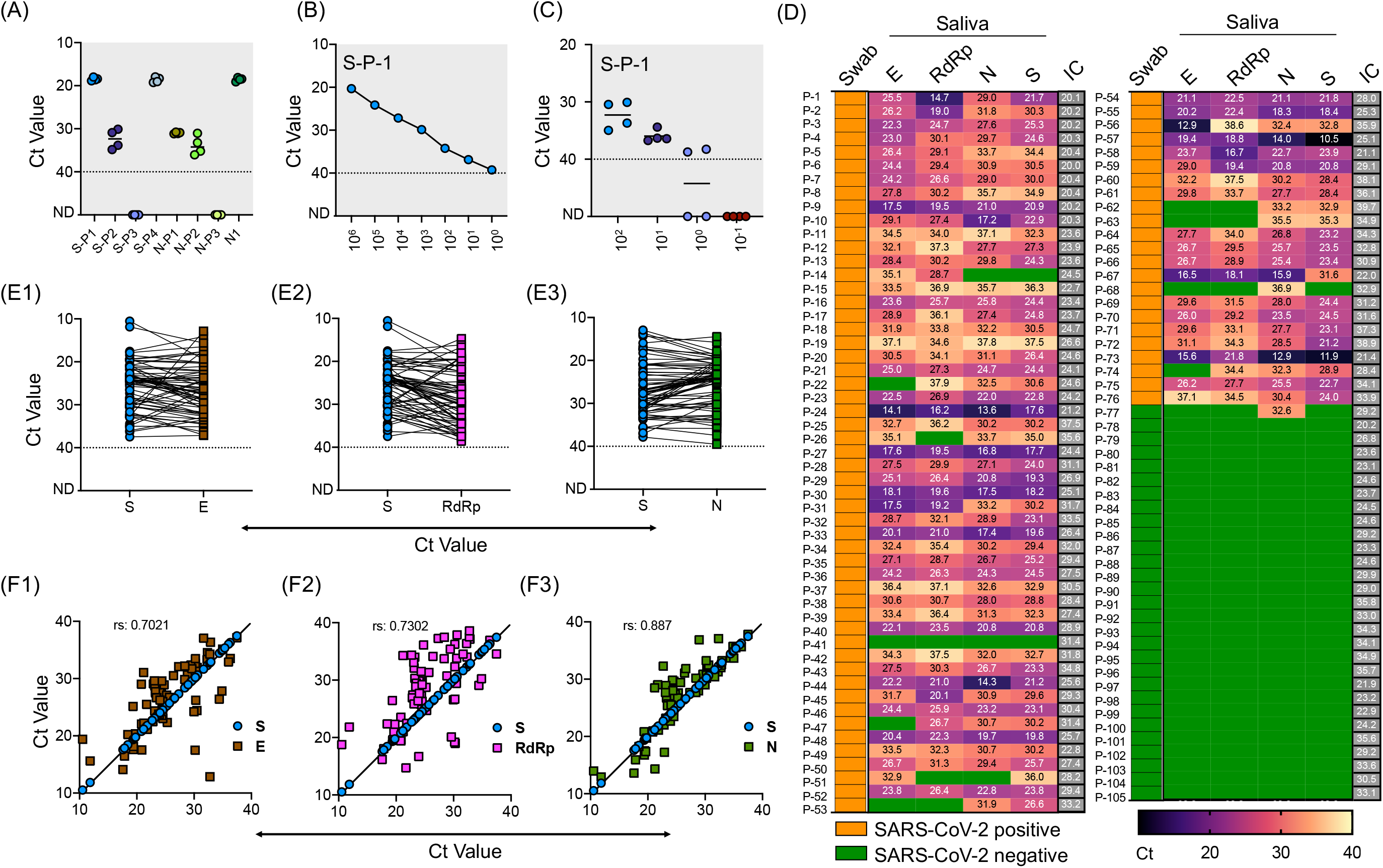
Validation of saliva-based detection of SARS-CoV-2 in clinical samples. **(A)** Standardization of SARS-CoV-2 specific primer pairs for S and N gene. Primers are labelled as S-P1 to SP-4 for S gene and N-P1 to N-P-3 for N gene, N1 represents the CDC approved primer for N gene. Dotted line indicates lower level of detection; ND indicates not detected **(B**) Determination of limit of detection using RNA of S spiked into RNase free water with serial dilutions. **(C)** RNA extracted from four normal saliva samples was used at various copy numbers to find any interference for detection of spiked-in SARS-CoV-2 S gene RNA. **(D)** Heat map of 105 saliva samples showing RT-qPCR results represented as Ct values for E, N, RdRp and S genes respectively. The test results of these samples were validated by the hospital using swab samples, and represented as Swab. IC is the internal control. Green boxes represent the samples with Ct values not detected and were labelled as SARS-CoV-2 negative, and yellow boxes indicate swab samples positive for SARS-CoV-2. (**E1-E3**) Shows the Ct value comparison between S gene with E, RdRp, and N gene respectively. (**F1-F3**) Shows the spearman correlation of Ct values between S gene with E, RdRp, and N gene respectively.

Similarly, we used RNA extracted from four saliva samples of the volunteers to find any cross-reactivity. These samples were collected in 2018 for an unrelated study and stored at −80°C. Similar amplification profile and LoD was obtained with spiked-in RNA in these samples, while lack of amplification without spiked-in RNA suggests that no interference and no-cross reactivity exists with the RNA from the saliva samples (Figure 1C). After finding LoD, we used these same sets of primer/probes along with N1, RdRp, and E gene and performed RT-qPCR in the clinical saliva samples using a commercially available kit.

We obtained 113 clinical saliva samples from patients with matched swabs tested at Safdarjung Hospital, New Delhi. Based on the swab results for which the hospital performed the RT-qPCR, 82 samples were positive, and 31 were negative. We extracted RNA from these samples using the viral RNA extraction kit (see Methods) followed by one-step RT-qPCR analysis using the commercially available kit and compared the results with our in-house optimized protocol. Based on previous reports and the LoD derived for S gene, we set the upper limit for detection at Ct < 40 to mark the sample as positive, and samples with Ct above 40 were marked negative. In the initial screening, 8 samples (6 positive and 2 negative) showed no detectable signal for internal control and hence were eliminated for the analysis. Thus, a total of 105 saliva samples (76 positive and 29 negative) were used to perform the RT-qPCR. We found 98.7% positive agreement with swab results with at least one of the primer/probe sets tested. Individually, we found 97.4% agreement for E and S gene, 89.6% for the E gene, and 90.9% for RdRp.

Among the negative samples, all the primer/probes for E, RdRp and S showed a 100% positive agreement, while one sample with a negative swab result also showed a positive signal for the N-1 gene primer/probe (Figure 1D). Comparative analysis of our in-house optimized protocol with a commercially available kit revealed a close correlation of S gene with N (r = 0.887) (Figure 1E3, 1F3), which was comparatively better than comparison for S gene with E gene (Figure 1E1, F1) and RdRp (Figure 1E2, F2). Thus, our in-house optimized RT-qPCR method is in high-agreement with the CDC-approved N-1 gene-based amplification. Further, these results confirm that saliva and swab samples have a high degree of consistency, and saliva can decisively detect SARS-CoV-2 in COVID-19 patients. Our results confirm the findings of the previously published reports which have demonstrated the use of saliva as a reliable clinical sample for detecting SARS-CoV-2 with a detection limit and sensitivity comparable with the nasopharyngeal and oropharyngeal swab (hereafter swab) (Fakheran et al., 2020; Procop et al., 2020). While some other reports have shown slightly better analytical sensitivity of saliva samples (0.98 virus RNA copies/ml) than other biological fluids for SARS-CoV-2 detection (Wyllie et al., 2020), we did not perform a direct comparison.

### RNA extraction-free detection of SARS-CoV-2 in clinical saliva samples

A major hurdle in COVID-19 testing is the need for viral RNA extraction, which poses a challenge to speed-up the testing. We envisioned to use a simple RNA extraction-free (hereafter, RNA_ExF) method and validate its analytical sensitivity on SARS-CoV-2 clinical samples. Recently, several RNA_ExF methods were employed to test their analytical sensitivity in clinical samples; however, these methods have their own benefits and limitations (Alcoba-Florez et al., 2020; Brown et al., 2020; Bruce et al., 2020; Grant et al., 2020; Hasan et al., 2020; Merindol et al., 2020; Srivatsan et al., 2020; Wee et al., 2020). Thus, we aimed to develop a more sensitive workflow for the RNA extraction-free testing of saliva samples. We initially optimized Proteinase K concentration under various heat inactivation conditions and tested by introducing 10^5^ copies of the S gene into the normal saliva. We found that at a concentration of 1.25 mg and dual heat inactivation (37 °C for 10 mins and 95°C for 5 min), a better analytical sensitivity could be obtained in comparison to the heating of samples at 65°C or with higher concentrations of Proteinase K (Figure 2A). While at all concentrations tested, the detection limit was relatively less than the samples in which the S gene was spiked into RNase free water. As saliva contains mucoproteins, which may interfere with the detection, we next tried mucoactive chemicals (sodium citrate and ammonium chloride) and mucolytic agent N acetylcysteine (NAC).

**Figure 2:**
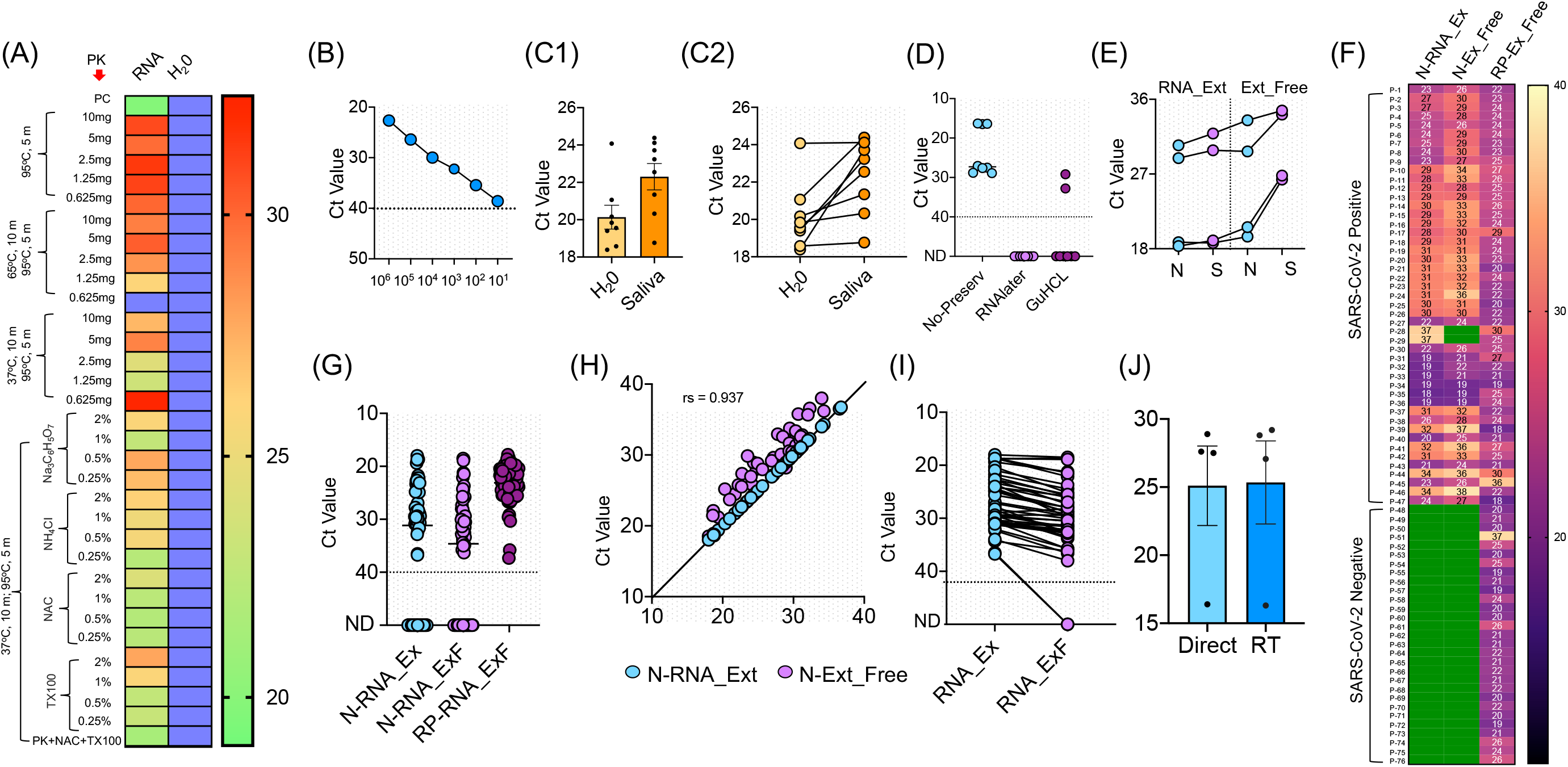
RNA extraction-free detection of SARS-CoV-2 in saliva samples. **(A)** Heat map of Ct values obtained with 10^5^ copies of S gene standard RNA spiked into normal saliva and subjected to various heat inactivation and chemical treatments. PK: Proteinase K; NAC: N-acetyl cysteine and TX100: Triton X100. Saliva samples with water was used instead of spiked-in RNA as negative control, which showed no detectable Ct value; blue boxes. **(B)** Standard curve showing various dilutions of S gene RNA spiked into normal saliva to obtain the LoD. **(C1, C2)** Comparison of Ct detected when S gene RNA was spiked into the saliva samples of 8 SARS-CoV-2 negative volunteers vs the same spiked into water as control. **(D)** Optimization of various RNA storage agents like RNAlater, guanidinium hydrochloride (GuHCL) and without RNA storage agent for detection of SARS-CoV-2 in 8 clinical saliva samples. (**E**) Comparison of N and S gene amplification in saliva samples after undergoing heat and chemical denaturation. **(F)** Heat map of Ct values obtained for N gene for 76 samples with RNA extraction (N-RNA_Ex) and RNA extraction-free (N-RNA_ExF) method. Human RNaseP (RP) was used as the experimental control to find the RNA integrity of the samples used. Green boxes represent the samples with not detected Ct values. **(G)** Shows the individual Ct values of N-RNA-Ex and N-RNA_ExF along with the RNaseP with dotted line indicating lower level of detection. **(H)** Correlation of Ct values between N-RNA_Ex and N-RNA_ExF method **(I)** Median of Ct values of two methods as indicated by solid lines. The dotted line represents lower Ct value below which samples were labelled as not detected (ND). **(J)** Comparison of Ct values obtained from saliva samples when stored at room temperature (RT) for 6 hours with same samples processed without storage (direct).

Interestingly, we found that samples treated with NAC exhibited relatively better detection than other mucoactive agents. We also used Triton X100 in the formulation and found that addition of this non-ionic detergent at concentrations of 0.25-2% did not interfere with detection, and hence may aide in release of viral RNA when applied to clinical samples (Figure 2A). Thus, we found an optimal heat inactivation method (37°C for 10 mins and 95°C for 5 mins) and chemical composition which consists of proteinase K (1.25 mg/ml), NAC (0.5%), and Triton X100 (0.5%), to provide a simple buffer formulation for saliva-based detection of SARS-CoV-2.

To obtain the LoD, we used spiked-in S standard RNA into the saliva samples and performed chemical treatment and heat inactivation. We could accurately detect as low as 10 copies of RNA/reaction, which though slightly lesser compared to a single copy detection when RNA was spiked-in water (Figure 2B). We further tested this buffer’s sensitivity and heat inactivation on 8 saliva samples collected from volunteers in 2018 for an unrelated study. In comparison to the RNA spiked in water, we could detect the amplification in all saliva samples, while a slight increase in Ct values was observed, with a difference in the mean Ct of 2.169 ± 0.9526 (Figure 2C1, 2C2). Collectively, these results indicate that an optimized buffer and heat inactivation conditions are suitable for RNA detection in saliva samples with an RNA_ExF free workflow.

Based on the results that RNAlater solution maintains the RNA quality and detection sensitivity in clinical samples (above, Figure 1D), we initially used RNAlater solution and guanidine hydrochloride (GuHCL) to collect the saliva samples for validation using RNA_ExF workflow. We gave 3 sets of tubes for sample collection; the set-I with RNAlater, set-II with GuHCL, and set-III without any solvent. A total of 8 samples collected from the same patients were obtained and used for the analysis. We found that both GuHCL and RNAlater inhibited the assay detection with our optimized chemical and heat treatment. Simultaneously, the direct use of the samples without preservatives showed better detection (Figure 2D). Thus, these results suggest that the collection of saliva directly into the collection tube without any chemical or RNA preservative is optimal for RNA_ExF detection of SARS-CoV-2.

Next, we used the same optimized protocol to validate the analytical sensitivity of this workflow on 83 additional clinical samples. Each collected sample was divided into two separate tubes, one containing the RNAlater solution, and in the other tube, saliva was collected without any solvent. The samples collected in RNAlater were subjected to RNA extraction using the kit-based method, while samples collected without any solvent underwent chemical treatment and heat inactivation and were directly used for RT-qPCR analysis. Initially, we tested N and S primer sensitivity with the RNA extraction-free method and performed the assay in four samples. A slightly better sensitivity was detected when we used N primer in samples with RNA extraction-free method (Figure 2E). This discrepancy in the sensitivity of these two genes could be due to small amplicon size of N (72bp) than S (112bp). So, we used N primer and the RNaseP (RP) for the subsequent screening of the clinical samples. We used RP for initial screening to qualify the sample for comparative analysis between the RNA extraction and the extraction-free method. Only those samples with a detectable Ct value for RP (76 out of 83 samples) were qualified and further used in this study.

As shown in Figure 2F, 76 samples of COVID-19 were tested with RNA extraction-based method, out of which 47 tested positive, while 29 showed no detectable signal and were marked negative. With our optimized RNA extraction-free workflow, 45 out of 47 samples showed positive and 29 out of 29 showed negative results, with an overall 95.7% agreement for positive test samples and 100% agreement for the negative samples (Figure 2F, G). The two samples that showed no detectable signal in the RNA extraction-free method had a comparatively high Ct value obtained with the RNA extraction method (Ct: 37) (Figure 2I). Though the Ct values were slightly higher with the RNA extraction-free method, with a difference between the means of the two methods at 2.545 ± 1.158. Correlation analysis of Ct values reveal high degree of correlation between RNA extraction-free workflow with the RNA extraction method (spearmen coefficient, rs=0.937) (Figure 2H). Overall, these results suggest that saliva can be directly used for the detection of SARS-CoV-2 without the need for costly and time-consuming RNA extraction steps. This simple extraction free workflow thus overcomes the time-consuming and expensive RNA extraction step for SARS-CoV-2 detection and hence will eliminate the supply chain constrain and need for laboratory set-up to perform the assay.

To determine the sample stability over time, we conducted the assay on 4 SARS-CoV-2 saliva samples stored at room temperature. We observed no significant difference in the analytic sensitivity of the samples when stored up to 6 hours (Figure 2J). Thus, these results suggest the feasibility of home collection of the saliva samples without technical assistance, cold storage, or viral transport medium.

### Cas13a based detection of SARS-CoV-2 in saliva samples

After validating RNA_ExF detection of SARS-CoV-2 in saliva samples, we next explored detection methods, which are (1) relatively instrument-free, (2) previously validated for SARS-CoV-2, and (3) exhibit sensitivity consistent with RT-qPCR. To meet this criterion, we found the SHERLOCK-based detection method to be the most appropriate, which was also recently approved by the US FDA. SHERLOCK relies upon the collateral activity of Cas13a to cleave the colorimetric or a fluorescent reporter once the target molecule is detected (Figure 3A). To use this method on our optimized workflow, we obtained commercially synthesized and previously verified crRNA sequences for the S gene spanning the region for which we validated the RT-qPCR assay (Table S1). Similarly, we also obtained crRNA corresponding to the Orf1ab gene (Zhang lab, MIT). Cas13a was isolated from an Addgene plasmid (#90097) which was a kind gift from the Zhang Lab, MIT. The plasmid was propagated and purified based on the published protocol (Patchsung et al., 2020). Using the methodology employed by Kellner et al., we first performed SHERLOCK on standard RNA corresponding to S and Orf1ab to validate this method (Kellner et al., 2019). In addition, we also obtained the recombinant Cas13a from the commercial source to confirm the results.

**Figure 3:**
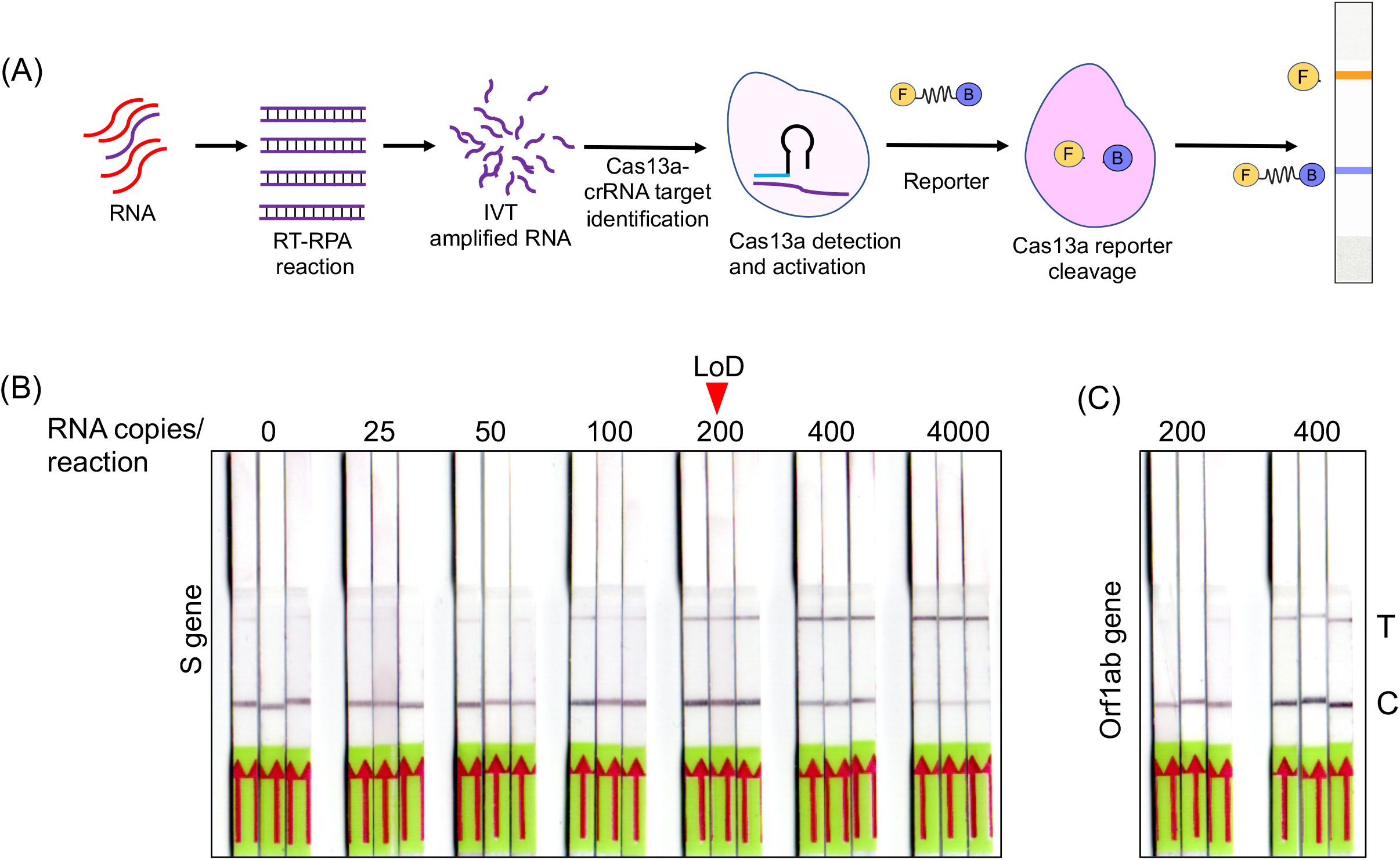
Optimization of SHERLOCK-based detection on extraction free saliva samples. **(A)** Schematic representation of various steps involved in SHERLOCK-based detection when the starting genetic material is RNA. Cas13a enzyme is used for the target recognition and reporter cleavage. For visual detection using LFA, RNA reporter molecule conjugated with 6-Carboxyfluorescein (FAM) and biotin is used. **(B)** Images of paper-strips after lateral-flow assay obtained from spiked-in saliva samples using S gene standard RNA with a range from 0 to 4, 000 copies of RNA/reaction. A consistent detection of test lane signal was obtained in all three samples with 200 copies of RNA, which was considered as LoD for visual readout. **(C)** Similarly, Orf1ab standard RNA was subjected to LFA and paper-strip images were obtained. The LoD for Orf1ab was found to be higher than S gene at 400 copies/reaction.

Previous studies have shown that both fluorescence-based detection and lateral-flow readouts using paper-strip can be accurately used to demonstrate the working principle of this method. Considering the ease-of-use, instrument-free detection, and cost-effectiveness, we choose the lateral-flow readout to validate this method and correlated it with the corresponding RT-qPCR data. A range of various dilutions of the standard RNA spiked-in control saliva was run, and the lateral-flow readouts were labeled as positive or negative by visual detection. As shown in Figure 3B, a consistent increase in the positive signal in the test lane was obtained, which corresponded to the lowest RNA copy number of 200 copies/reaction for the S gene with 100% detection sensitivity with a corresponding Ct value of 35.4. In comparison, the detection limit for Orf1ab was lower at 400 copies/reaction (Figure 3C). The LoD for the S gene was slightly higher than the spiked-in RNA samples, which is consistent with the results from other groups (100 copies/reaction for the S gene) (Joung et al., 2020). This discrepancy is probably due to the sample processing and assay for detection. While the study by Joung et al. used spiked-in RNA samples followed by RNA extraction, and Cas12-based detection; we used RNA_ExF samples and employed Cas13-based detection. Overall, these results confirm the previous reports which show that SHERLOCK-based approach integrated with visual lateral-flow readout can be used for SARS-CoV-2 detection.

### Validation of SHERLOCK-based detection on RNA extraction-free saliva samples

To make this tool affordable and accessible with a provision for field testing, we next performed SHERLOCK on the optimized RNA_ExF saliva samples. We divided the samples into 4 groups based on the Ct values. Group I with Ct > 25; group II with Ct between 26-30; group III with Ct between 31-35, and group IV with Ct < 36, with 5 samples in each group. Corroborating the RT-qPCR, 39 out of 40 positive sample also showed a positive signal with an LFA readout with Ct values below 35 (Figure 4A and Supplementary Figure 2A). Thus, SHERLOCK was in 98% agreement with the Ct values below 35, and by combining this method with RNA extraction-free saliva samples, we could obtain comparable sensitivity to the RNA extraction method for SARS-CoV-2 detection, as reported by others (Patchsung et al., 2020). To further validate the reliability of this approach, we performed a longitudinal detection analysis of the clinical samples. For this, we selected two patients who have been four times sampled (both swab and saliva) at the hospital at different time intervals after the symptom onset. We used the saliva samples and performed SHERLOCK assay, followed by lateral flow readout of the test results. We found that the SHERLOCK-based detection accurately confirmed the findings of the RT-qPCR swab results (performed in the hospital) in these test samples (Supplementary Figure 2B1, 2B2). Taken together, these results confirm that SHERLOCK-based diagnosis provides a reliable and extensive instrument free method for SARS-CoV-2, without compromising the assay’s analytical sensitivity.

**Figure 4:**
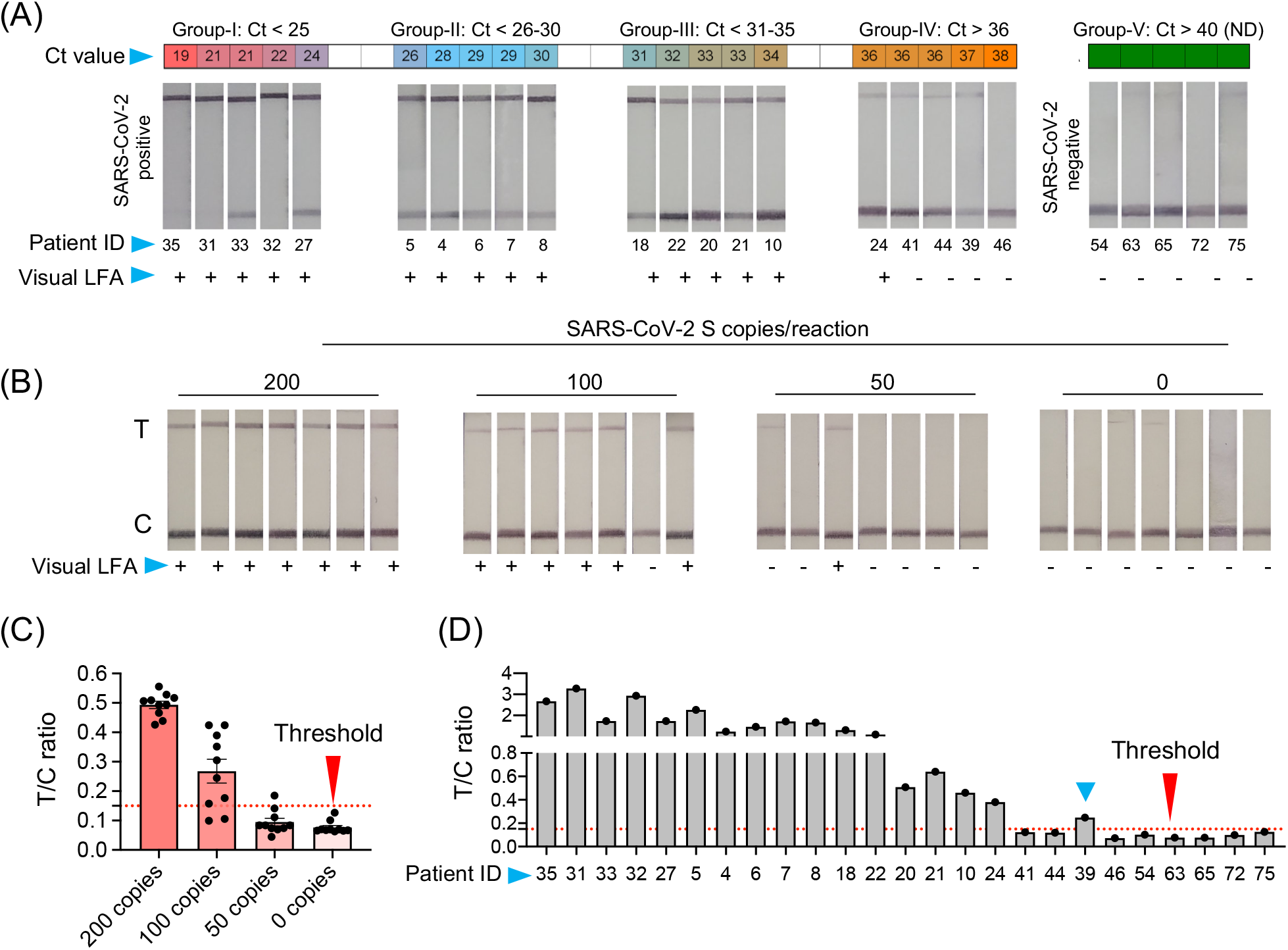
Validation of SHERLOCK on RNA extraction-free saliva samples. **(A)** RNA extraction-free samples were used for the detection with SHERLOCK-based method using visual lateral-flow. Samples were divided into five groups, based on the Ct values with Group-I Ct below 25; Group-II Ct between 26-30, Group-III Ct between 31-35, Group-IV Ct above 36, and Group-V Ct not detected (ND). The LFA images of samples with respective Ct values is shown along with the patient ID corresponding to samples in *Figure 2*. (**B**) Representative images of the 7 paper-strips with 200, 100, 50, and 0 RNA copies spiked-in to the saliva samples and subjected to SHERLOCK. **(C)** To obtain semi-quantitative analysis of the data, 10 images of paper-strips corresponding to 200, 100, 50, and 0 RNA copies as shown in *Figure 3b* and *Figure 4b*, were subjected to image quantitation. The threshold value (T/C ratio) was obtained based on the signal in the control (C) and test (T) lane. The T/C ratio was considered positive above the background value of 0.15. Based on T/C ratio, the detection sensitivity was found to be 80% with100 copies of RNA and 100% above 200 copies of RNA per reaction. (**D**) Similarly, visual results of paper-strips shown in Figure 4A, were subjected to signal quantitation and T/C ratio was calculated. One sample (blue arrow head, ID: 39), which was difficult to characterize by visual detection, was correctly characterized as positive using T/C ratio (blue arrow head).

### Semi-quantitative analysis of the LFA signal with provision for smartphone application

The limitation with lateral-flow readout using visual detection is that sometimes a weak signal may appear in the test lane. This usually occurs if too little or too much (High Dose Hook Effect) of the reporter molecule is used. Thus, a very precise amount of the reporter has to be added, which otherwise may interfere with test results. However, under laboratory free settings such as POCT and for home testing, the precision in handling may be challenging and it is expected that a background signal may appear in the test lane, which may sometimes be difficult to interpret by naked eye. Further, based on our results and previously published studies (Patchsung et al., 2020), SHERLOCK with visual readout performs exceedingly well with samples at a higher copy number of the analyte. Though, the interpretation of results become difficult when samples with very low copies of analytes are present, where the signal in the test lane may be close to the background, which besides the above-mentioned issues may also in some instances appear due to non-specific probe degradation (Patchsung et al., 2020). Thus, to determine the background signal and differentiate between the noise and actual signal, we used ten replicates of negative control and samples with 50, 100, and 200 copies of RNA/reaction spiked into normal saliva and subjected to chemical treatment and heat inactivation, respectively (Figure 4B). The SHERLOCK assay was done and the paper-strip images were obtained and processed using Image J (see Methods). A threshold value was generated based on the ratio of signal intensity in test lane to control lane (T/C) corresponding to no RNA and three known concentrations of standard RNA. Based on the signal obtained from negative samples, we obtained a threshold value with a T/C ratio of 0.15, and below this ratio, the samples were labeled as negative (Figure 4C). By quantifying the signal of various dilutions, all samples with 200 copies of RNA showed T/C ratio above threshold (positive). In samples with 100 copies, nine showed positive T/C ratio, though most of these sample results were difficult to interpret by visual detection. Thus, we set this new LoD to 100 copies based on signal quantitation, which corresponded to a Ct value of about 36.32 (Figure 4C).

Next, we quantified the signal from the respective positive and negative clinical samples. A 100% agreement was observed in the negative samples between visual and T/C ratio. Similarly, samples with Ct > 35 also showed 100% agreement. Surprisingly, among five samples with Ct < 36, two showed T/C signal above threshold, in contrast to one sample detected by the visual readout (Figure 4D). Thus, while visual detection accurately detects the signal below a Ct value of 35, it suffers the detection limit in samples with very low viral RNA copy numbers. Under such circumstances, a semi-quantitative approach is more feasible, as quantitation is provided based on the ratio between the test and the control lane instead of solely relying on the signal at the test lane. Overall, these results suggest that LFA signals obtained using RNA_ExF saliva samples can be precisely quantified and correlated with Ct values to give a fair estimate of the viral load (at higher Ct) and further enhance the analytical sensitivity.

To further ease the interpretation of test results, we developed mobile application and integrated it with the lateral-flow test strips (Figure 5, Video 1 & 2). The smartphone application has also provision to save the test results which can be easily accessed for research purpose. Further, the application has an online consultation option, which the patients can access to get immediate help related in performing the assay – an important development when performing home testing. The mobile application can be downloaded using the link given in the methods.

**Figure 5:**
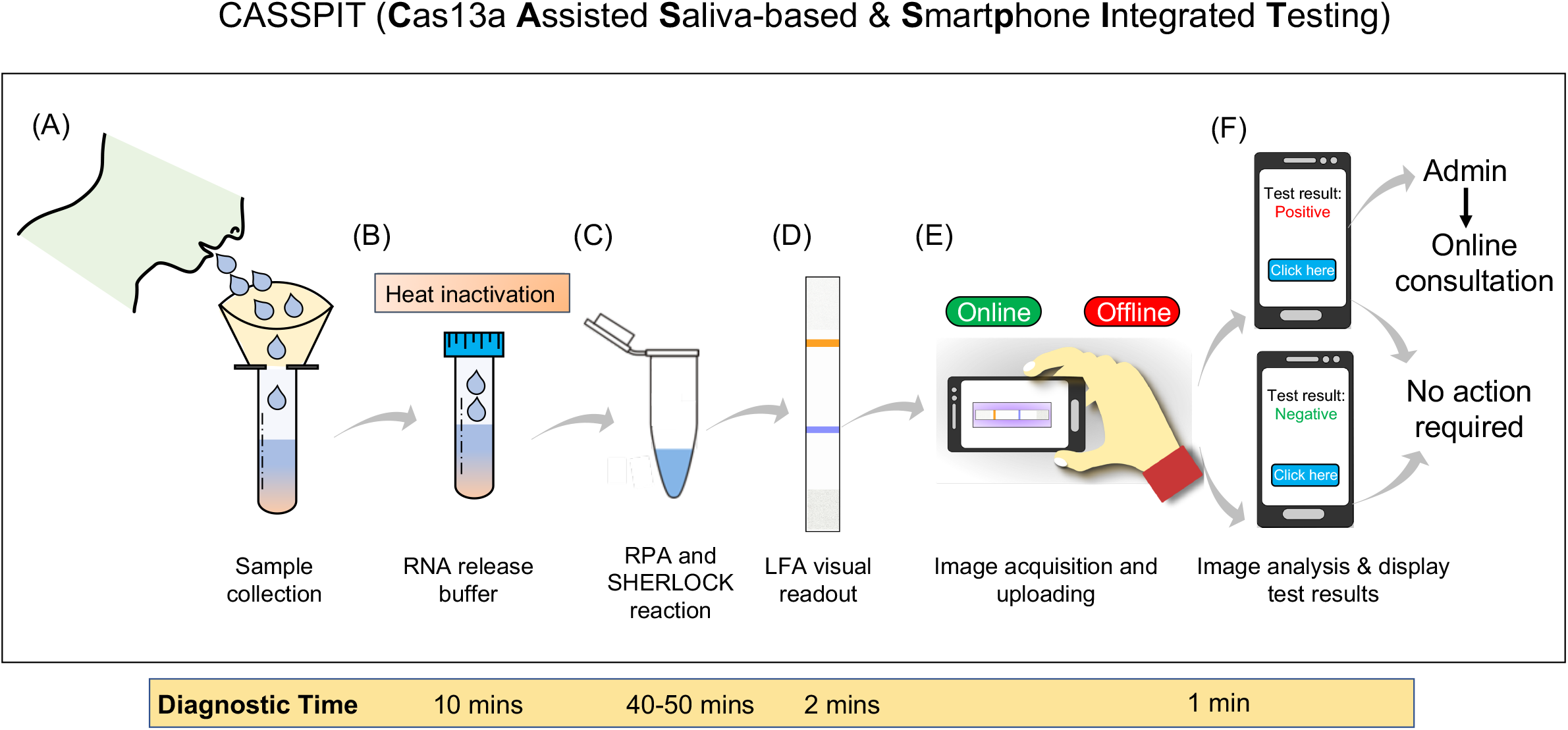
Schematic representation of the CASSPIT workflow. **(A)** Self-collection of saliva samples by the patient in a sample collection tube which contains the RNA release chemical agents. **(B)** The samples will be subjected to heat inactivation to inactivate the virus and simultaneously release the viral RNA. **(C)** Released RNA will be transferred into one-pot or two-pot SHERLOCK master mix tube to amplify the signal with RT-RPA and detect the target by Cas13a. After target detection, activated Cas13 will cleave the reporter. **(D)** Paper-strips will be immersed in the SHERLOCK reaction mix and subjected to LFA to obtain the visual results. **(E)** Using mobile phone camera, images of the paper-strip will be captured and subjected to processed using the app. Test results will be provided based on the signal detection in Test and Control lanes of the paper-strip. Further the app will have a provision to store the images, test results and help with online assistance if needed. The overall workflow with saliva as test samples is named as **C**as13 **A**ssisted **S**aliva-based & **S**mart**p**hone **I**ntegrated **T**esting (CASSPIT).

## Discussion

Considering the impact of COVID-19 on global health and economy, it is imperative to have at our disposal field-deployable diagnostic test kits that are robust, cost-effective, specific, and sensitive. The onset of COVID-19 pandemic has witnessed an upsurge in molecular diagnostics of SARS-CoV-2, but only a few of them have sensitivity and specificity comparable to the gold standard RT-qPCR, especially those based on nucleic acid detection such as CRISPR-Cas system (Broughton et al., 2020; Patchsung et al., 2020). However, the major challenge to use these tools as a POC device for field testing or home testing persists. In addition, workflow of these methods is contingent on additional RNA extraction steps that require trained professionals and sophisticated laboratory set-up and instrumentation to perform the assay. Considering these fundamental challenges, we developed, optimized, and validated the use of a simple workflow to detect SARS-CoV-2 in saliva samples for the following reasons: (1) saliva can be self-collected with ease to minimize direct contact with the healthcare workers, and to reduce handling errors; (2) repeated sampling is feasible without incurring discomfort to the patient, with a reasonably uniform sample distribution; and (3) higher stability of the SARS-CoV-2 in saliva even when stored at room temperature (up to 7 days) (Vogels et al., 2020b).

Initially, we validated the analytical sensitivity of clinical saliva samples following RNA-extraction and RT-qPCR assay for detection, and found a close agreement with corresponding swab test results performed by the hospital. Next, we developed a simple workflow to detect SARS-CoV-2 in saliva samples without need for an extra and time-consuming RNA-extraction step. Optimization of this workflow was challenging as unlike other biological fluids, the molecular composition of saliva hinders RNA detection, besides being more amenable to RNases (Ochert et al., 1994; Ostheim et al., 2020). Also, SARS-CoV-2 being an enveloped virus, the RNA release condition had to be optimized so that its degradation is minimized. Working on various chemical treatments and heat inactivation steps, we formulated a unique buffer composition containing the optimal concentration of Proteinase K, Triton X 100, and N-acetyl cysteine, followed by heat inactivation. Together this buffer and heat condition were sufficient to cause the release SARS-CoV-2 RNA and maintain its stability. On testing this workflow on clinical samples, we found a close agreement with the kit-based RNA extraction method, which is used for detection of SARS-CoV-2. Our results were thus consistent with previous reports on the use of RNA extraction-free detection of SARS-CoV-2 (Alcoba-Florez et al., 2020; Brown et al., 2020; Bruce et al., 2020; Grant et al., 2020; Hasan et al., 2020; Merindol et al., 2020; Srivatsan et al., 2020; Vogels et al., 2020b; Wee et al., 2020).

To further simplify the diagnostic workflow, we have optimized the RNA extraction-free detection of SARS-CoV-2 with two-(step)pot SHERLOCK, which relies on the collateral activity of Cas13a – recently validated on clinical COVID-19 samples (Hou et al., 2020; Patchsung et al., 2020). Our results indicate that the two-pot SHERLOCK and RT-RPA reaction works exceedingly well in saliva samples with RNA extraction-free method, and found 98% positive agreement with the RT-qPCR data, with Ct>35, corresponding to approximately 200 copies/reaction. In view of the reports that most SARS-CoV-2 patients have a cut off Ct values around this range (33.5 for E; 33.5 for RdRp; and 34.5 for N gene) (Uhm et al., 2020), our approach can be reliably used as an alternate to RT-qPCR with provision for RNA_ExF free workflow. The LoD which we obtained for extraction free saliva samples with a two-pot reaction is slightly lower than what others found when RNA-extraction methods were used (Patchsung et al., 2020). This could be due to: (1) High DNA, proteins, and other ions present in saliva, which interferes with the detection, irrespective of the type of detection method used, i.e., RT-qPCR or SHERLOCK (Ochert et al., 1994; Ostheim et al., 2020). (2) Ambiguity in visual detection, when the test lane signal is shallow and difficult to differentiate by the naked eye.

Thus, to further improve the detection sensitivity of these test results, we employed image-based signal quantification of lateral-flow strips. With this approach, we obtained a T/C threshold ratio, which differentiated between the positive and negative samples more precisely than visual readout, and obtained an improved LoD of 100 copies of RNA per reaction. This improved LoD based on T/C ratio is similar to the LoD obtained by Joung et al. (100 copies/reaction) with one-pot Cas12a and RT-LAMP reaction (Joung et al., 2020). Other groups have attained slightly better sensitivity with two-pot reaction. Using Cas12a and RT-LAMP, Broughton et al. achieved an LoD of 24 copies/reaction with lateral-flow readout on SARS-CoV-2 clinical samples (Broughton et al., 2020). Similarly, using Cas13a and RT-RPA, Patchsung et al. achieved an LoD of 42 copies/reaction in spiked-in saliva or nasopharyngeal samples (Patchsung et al., 2020). Thus, these studies indicate that there is scope to further improve the analytical sensitivity of RNA_ExF workflow.

Another disadvantage of paper-strip-based methods is that these test results are difficult to access for clinical studies. That includes data for survey, vaccine trials, or testing other therapeutic interventions. To overcome this challenge, we integrated the lateral-flow readout with a mobile application with provision for offline or online mode. The smartphone integrated workflow will thus provide an accurate estimate of the signal in test vs control lane, that will remove any ambiguity associated with visual detection. Further, this application will provide access to image files, patient clinical parameters, and LFA quantitative results. A video demonstration of a saliva-based workflow integrated with the mobile application is shown (Video 1 & 2).

In summary, we provide a simple workflow for SARS-CoV-2 detection, which is a unique addition to the rapid, cost-effective, and straightforward diagnostic methods. Such user-friendly testing methods have an immediate application under the settings where trained professionals and costly instruments are limited. Further, owing to the high-sensitivity and high-specificity of Cas13a, our optimized workflow on saliva samples could provide a rapid and better alternative to the existing detection methods and speed up the testing. We envision that in coming time, CRISPR-Diagnostics based on either Cas13a or a similar method which utilizes Cas12 (Broughton et al., 2020; Nguyen et al., 2020) and integrated with a smartphone-based readout (Ning et al., 2020) will be a better alternative to RT-qPCR based testing under resource limiting settings. Moreover, integrating CRISPR-Diagnostics with RNA extraction-free workflow and smartphone application will provide an alternative to error-prone rapid antigen tests to scale-up COVID-19 testing across resource-constrained areas and intensify trace, test, and treat strategy for COVID-19.

The future of CRISPR-Diagnostics is promising as it provides rapid, accurate, low cost, and laboratory free genetic testing. Our study has shown feasibility of this diagnostic approach with RNA extraction-free saliva samples which can be extended to home testing. There is still scope to further improve these tools, especially for use in fully instrument free settings and without pre-amplification step. While a recent study has provided an improved version of CRISPR-Diagnostics, which bypasses the amplification step (Fozouni et al., 2020), more optimization is needed to provide a fully instrument-free diagnostic platform. Importantly, CRISPR-Diagnostics has the provision to detect single nucleotide mismatches and hence will serve as a rapid diagnostic tool to detect any new mutations arising in the SARS-CoV-2 genome (Kirby T, 2021). Some of these mutations have already been shown spreading at a higher rate and thus it is imperative to adopt and deploy CRISPR-based diagnostics to extensively screen the transmission of these new SARS-CoV-2 strains.

## Methods

### Patient information and ethical statement

The work was indented to develop a simple workflow for SARS-CoV-2 detection. The clinical samples collected for the work were used after obtaining Institutional ethical clearance from the Safdarjung Hospital (IEC/VMMC/SJH/Project/2020-07/CC-06). The ethical clearance was also obtained from Jamia Millia Islamia (1/10/290/JMI/IEC/2020). Further, biosafety clearance was obtained from Jamia Millia Islamia (Ref.No.P1/12-21.12.2020). Patient consent was obtained to collect the samples according to the ICMR GCP guidelines.

### Sample collection

A total of 210 clinical saliva samples were collected from Safdarjung hospital from June till November 2020, New Delhi. The saliva samples were collected from the patients at the same time when swab was collected for COVID-19 testing by the hospital. The hospital provided the swab RT-qPCR confirmatory results of 201 samples, while 9 samples had no corresponding confirmatory test results done in the hospital and were labelled as blind samples. Initially all the samples were collected in RNAlater solution for validation of the saliva-based detection of SARS-Cov-2 with RT-qPCR. Subsequently, samples were divided into 2 parts; (part 1) was collected in RNAlater solution, and (part 2) was collected in tubes containing proteinase K (1.25mg), Triton X 100 (0.5%), and NAC (0.5%). Similarly, samples for longitudinal studies were also collected from two patents (n=4 each). All the samples were processed in NABL certified (MC-3486) and ICMR approved Diagnostic laboratory for COVID-19 testing following the regulatory guidelines and protocol (360 Diagnostic and Health Services, Noida, U.P., India).

Samples were collected at different time interval in the hospital and stored at −20°C until carried to the facility for further processing. The time between sample collection to processing was around 3-5 days.

### Plasmids, primers and synthetic DNA fragments

Plasmids corresponding to S and N genes were received as a gift from Krogan laboratory, Department of Cellular and Molecular Pharmacology (San Francisco, CA 94158, USA). These same plasmids are now available with Addgene (#141382 for S gene and #141391 for N gene). After propagating, plasmid DNA was isolated using commercially available DNA isolation kit (Vivantis technologies). For RT-qPCR, 1.0 ng of the DNA was used for respective genes and a set of 8 primers were optimized. These primers were synthesised in-house using online primer design tools or obtained based on previously validated sequences such as N-1 primers. The list of the primers is given in the Table S1.

Synthetic gene fragments for S gene and Orf1ab corresponding to the sequences given in the Table S1, were obtained from Xcelris Genomics, India and Bioserve biotechnologies, India. The gene fragments were synthesised along with the T7 promoter sequence.

### Viral RNA release from saliva samples

Various heat-inactivation steps, chemical components and buffers were used to find the optimal assay condition for the detection of the viral RNA. Heat inactivation optimization was done at 37°C for 10 mins and 95°C for 5 mins. Non-ionic detergents like Triton X 100 (Sigma) and Proteinase K (Sigma, Vivantas, and promega) were used at various combinations to find the optimal reaction composition. Optimum detection of spiked-in S gene was obtained at a concentration of 1.25 mg/ml for proteinase K and 0.5% of Triton X100 with either two step heat-inactivation (65°C or 37 °C for10 mins and 95°C for 5 mins) as well as with a single heat-inactivation step (RT for 15 mins and 95°C for 5 mins). Further, in order to minimize the interference of the mucoprotein in saliva, mucoactive agents like sodium citrate (Sigma) and ammonium chloride (Sigma); and mucolytic agent N-acetyl cysteine (Sigma) were used at various concentrations.

### RNA extraction and RT-qPCR

RNA extraction was performed as recommended (Qiagen viral RNA extraction kit). 140 µl sample was processed according to the protocol as per manufacturer’s instruction. Final elution of the RNA was done in 30ul of the elution buffer and 2 µl of the extracted RNA/reaction was used in one-step RT-qPCR analysis using the commercially available RT-qPCR kit for SARS-CoV-2 which contains there targeting genes E, N and RdRp along with the internal control. Similarly, we also used our in-house optimized primers/probes for validation. For RNA_ExF saliva samples, we used 4 µl of the input saliva sample per reaction. RT-qPCR was performed on Rotor gene Q (Qiagen) with the recommended reaction condition for the commercial kit (Allplex, Seegene). For S-P-1 and S-P-4, the following RT-qPCR conditions were used: Initial denaturation 95ºC for 5 mins, second cycle of the reaction include denaturation at 95ºC for 30 sec, annealing at 62ºC for 30 sec, and extension at 72ºC for 30 sec for 40 cycles.

### Synthetic gene block and T7 reverse transcription

To determine the limit of detection (LoD), we generated synthetic gene fragments for S and Orf1ab corresponding to the regions for which crRNA has been previously validated (Zhang Lab protocol, MIT and Table S1). Both single stranded gene fragments were obtained commercially (Xceliris Genomics). Each (1ng/µl) oligonucleotide fragments were first converted into double strand by end point PCR then purified (0.5 µg/µl) of the double stranded DNA fragment was used as a template for invitro transcription reaction using in vitro transcription kit (T7 Ribomax, promega, cat no-P1320). The invitro transcribed RNA oligonucleotide fragment was then purified using RNA cleanup kit (Vivantis, cat no-GF TR 050) and eluted in a final volume of 50 µl. The purified RNA of the respective gene fragments was used as the standard to determine LoD. Various dilutions of S gene standard RNA were made in nuclease free water corresponding to 10^0^ to 10^6^ copies/µl. Similarly, the standard RNA (10^5^ copies/µl) was spiked into the saliva samples for optimizing various chemical and heat-inactivation conditions.

### Expression and purification of cas13a protein

For the expression of Cas13a protein, the pC013-TwinstrepSUMO-huLwCas13a plasmid was transformed into E. coli BL21 (DE3) cells. The transformed cells were grown overnight in 10 ml LB media containing 100 µg/ml ampicillin antibiotic at 37ºC/180 rpm in incubator shaker. Next day, 5 ml of the overnight grown culture was inoculated in 1liter LB media containing 100 µg/ml Ampicillin. After reaching the growth of the culture to OD between 0.4-0.6, the culture was incubated at 4ºC for 30 mins. Before induction of the protein, 1ml of culture was taken for SDS-PAGE analysis. Expression of the protein was induced by adding 1 ml/0.5 M IPTG and 2% glycerol to pre-chilled culture and incubated in cooling (21ºC) incubator shaker for 16 hours at 300 rpm. After that, the culture was harvested by centrifugation at 6000 rpm for 10 mins at 4ºC. supernatant was discarded and pellet was lysed in the lysis buffer.

### Lysis of the cell pellet

The cell pellet was resuspended in lysis buffer (20 mM Tris-HCl pH-8.0, 500 mM NaCl, 1 mM DTT, 1x protease inhibitor cocktail sigma, and 0.5 mg/ml lysozyme). The cell resuspension was lysed by sonication (Sartorius Stedim) using 50% pulse amplitude (on 10 sec and off 20 sec) until completely lysed. The lysate was centrifuged at 12,000 rpm for 30 mins at 4 °C. The protein was then applied to a HiTrap SP HP column equilibrated with equilibration buffer (20 mM Tris-HCl pH 8.0, 1 mM DTT, 500 mM NaCl, 15mM imidazole). The supernatant fraction was passed five times from the column for complete binding of the protein. After washing with the binding buffer to remove nonspecific binders, the recombinant His6–SUMO–LwaCas13a was eluted in a linear gradient (with increasing the concentration of the imidazole from 20-500mM) of elution buffer. The best elution was obtained at 100 mM imidazole. Elution was done in the volume of 5 ml. For the cleavage of His tag, the eluted fraction was supplemented with 20 µl of sumoprotease (invitrogen #125880-18,1U/µl) and 7.5 µl of NP-40. The reaction mixture was added to the column and incubated at 4°C for overnight with gentle shaking. Next day, cleaved native LwaCas13a protein was obtained by draining the column. After draining of the column, it was washed with elution buffer containing 500 mM imidazole to ensure the complete cleavage of His-tag from LwaCas13a protein.

The drained native protein was subjected to concentration to 0.25 ml using centrifugal spin filter (50 MWCO-MERCK millipore) at 4000 rpm for 15 mins at 4°C. Then 5 ml of protein storage buffer (1M Tris-HCl pH-7.5, 5M NaCl, 5% glycerol and 10 µl DTT) was added to the same filter and again centrifuged at the same condition. Storage buffer containing native Lw-Cas13a protein was diluted to 2 mg/ml in storage buffer and stored at −20°C, as described previously (Kellner et al., 2019).

### RT-RPA and SHERLOCK assay

As described previously (Gootenberg et al., 2017; Kellner et al., 2019), RT-RPA was performed using the commercially available RPA kit (TwistDx). First, each RPA tube was divided into four reaction tubes by diluting the lyophilized mix in the RPA buffer (40 µl). For LoD, forward and reverse RT-RPA primers were added 1µl (10 µM stock) to each reaction tube along with 1 µl of reverse transcriptase enzyme (EpiScript). 4 µl of various dilutions of the standard RNA for S and Orf1ab were used as template (0 to 4000 copies/reaction). For RNA extraction-free samples, a total of 8 µl of sample input was used and the reaction components were adjusted accordingly. The reaction was initiated by adding 0.7 µl of magnesium acetate (280 mM stock). All the reagents were prepared and mixed at 4°C. Finally, the reaction mix was incubated at 42°C for 25 mins and tapped in between after every 3-5 mins. Particularly, tapping of the samples was found to exhibit better results than without and is highly recommended henceforth.

SHERLOCK assay of the above reaction mix was performed in a separate 1.5ml Eppendorf tube which contains 1 µl of the Cas13a either from extracted pool or commercially (MCLAB, USA, Cat no. CAS13a-200) obtained source (at 63.3 µg/ml concentration), 1 µl RNase inhibitor (20 U per µl stock; invitrogen), 0.6 µl T7 RNA polymerase (50 U per µl stock; Lucigen), 1 µl of crRNA for the respective genes (Synthego), 1µl of MgCl2 (120mM), 0.8µl of rNTP mix (100mM), 2ul of cleavage Buffer (400mM Tris pH 7.4), and 1µl (20 µM) reporter. 6 µl of the RPA mix from RNA extracted samples were used per reaction. The final volume of the reaction mix was adjusted to 20 µl with RNase free water. The reaction mix was incubated at 37 °C for 20-25 mins.

### Lateral-flow assay detection

The SHERLOCK reaction mix was subjected to lateral-flow assay using the commercially available test trips and buffer (Millenia Biotec). To the above reaction mix 80 µl buffer (HybriDetect assay) was added, provided with the kit. The visual readout of the test results were obtained by dipping the test strips (Milenia Biotech1T) into the respective 1.5 ml Eppendorf tubes and the reaction mix was allowed to flow for 2 mins.

### Analysis of the Lateral-flow signal to provide a semi-quantitative estimate of the results

To provide a semi-quantitative analysis of the lateral-flow readout, we used Fiji image J software to analyze the signal in the respective T and C bands. The corresponding band intensity of the test lane (T) and control lane (C) were calculated using integrated density parameter. The image of each strip was captured using a mobile phone (width=50, height=220). For the quantification, the image was further cropped to image size 40by220 which thus removed the outliers from the image. Any background noise from the image was subtracted using the rolling ball background subtraction method by keeping radius=50. All the images (40by220) were further thresholded by applying lower threshold value 0 and the upper threshold values between 240-245. Finally, the single band was segmented from each image in the frame size of 30by30 and integrated density was analyzed for the respective bands. The threshold value of T/C was calculated and found to be 0.15 above which the samples could be labelled as positive.

### Smartphone application for lateral-flow test results

The mobile application for detection and quantification of lateral-flow strips was developed using machine learning tools. The algorithm was implemented using the OpenCV package v.4.3.0 in Python 3.7.3. The Android App was developed with Android Studio v4.2 RC 2 (Google) with Java 8 and Gradle v4.1.0. To provide a clean user interface, the main screen was limited to a “COVID-19 Test” bottom tab button that opens up an in-app camera view to capture the image followed by custom image edit options. The image acquisition is only allowed through the mobile application to accurate documentation of taken images and test results. The image captured in the application is obtained as a Uri object, that is used for conversion to a byte’s array. In order to obtain the image analysis outcome, the bytes array obtained is passed as an argument to the Python backend script running through Chaquopy v6.3.0 that is a software development kit used in Android development environment. The bytes array image is further processed by the integrated image analysis module of the application. In the first image pre-processing step, the acquired image is converted to grayscale and region of interest is localized in the image through adaptive thresholding and edge detection techniques. In the next step, the perspective and orientation corrections are performed on the image which are present due to un-controlled mobile imaging. The shape of the strip and marker are two primary cues used for the aforementioned corrections. The resultant image is centered around the localize region of interest and cropped to obtained the strip area image to a fixed scale. The sample and control band image regions are detected from the cropped image based on the known strip structure and darker intensity profile of the bands. The mean intensity of each band region is obtained and ratio of the sample to control band in then calculated and displayed on the mobile application to the user along with the cropped strip image. The mean intensity of the sample band region is scaled to the range of 0-1 w.r.t a pre-defined reference point in situation when control band has weak visual appearance (signature).

The mobile application can be downloaded using the link: https://bit.ly/research-app.

### Statistical analysis

Statistical analysis of the samples was performed using GraphPad8 Prism. Spearman correlation coefficient was used for correlation analysis. Non-parametric t test was used to compare the mean difference between two data sets as mentioned in the results.

## Data Availability

We have followed all the necessary guidelines and archived the data related to this study.
Supplementary data files are separately uploaded and original data files are documented as per the guidelines.

## Abbreviations

CDC: Centers for Disease Control
COVID-19: Coronavirus Disease 2019
CRISPR: Clustered regularly Interspaced Short Palindromic Repeats
DETECTR: DNA Endonuclease Targeted CRISPR Trans Reporter
LAMP: Loop-mediated isothermal amplification
LFA: Lateral Flow Assay
NAC: N-acetyl Cysteine
PCR: Polymerase Chain Reaction
PK: Proteinase K
POC: Point of Care
RPA: Recombinase Polymerase Amplification
RT-LAMP: Reverse Transcription Recombinase Polymerase Amplification
RT-RPA: Reverse Transcription Loop-mediated isothermal amplification
RT-qPCR: quantitative Reverse Transcription Polymerase Chain Reaction
SARS-CoV-2: Severe acute respiratory syndrome coronavirus 2
SHERLOCK: Specific High-sensitivity Enzymatic Reporter un-LOCKing
US FDA: United States Food and Drug Administration

## Acknowledgement

Dr. Tanveer Ahmad is thankful for UGC for the start-up grant support (F.4-5/2018 FRP Start-up grant). Dr. Mohan C Joshi is thankful for DBT/Wellcome Trust India Alliance grant number IA/I/15/2/502086 the support. Dr. Jawed Iqbal is supported by the Ramalingaswami Fellowship grant (BT/RLF/Re-entry/09/2015) from Department of Biotechnology (DBT), and Early Career Research Award grant (File No. ECR/ 2018/002114) from Science and Engineering Research Board (SERB), Department of Science & Technology, Government of India. Iqbal Azmi and Md Imam Faizan acknowledge ICMR for Senior Research Fellowship. We are thankful to Dr. Arpita Rai for providing us the control saliva samples and all other faculty members of MCARS for their valuable support and discussion while preparing this manuscript. We thank Dr. Manish Kumar, BLDE university, Karnataka, for suggestions during the optimization of saliva-based workflow. We are thankful to Dr. Sushant G Gosh and Syed Kazim Naqvi for their support in establishing the molecular diagnostics facility at MCARS.

## Authors Contribution

TA, MJ, and JI designed the experiments and provided financial support for the research. IA, MF, TA, DKS and NC performed experiments. RK and SY collected clinical samples. RB and GDJ facilitated experimental work. AG and GJ designed mobile app.

All authors contributed to the article and approved the submitted version.

## References

Alcoba-Florez, J., González-Montelongo, R., Íñigo-Campos, A., De Artola, D.G.-M., Gil-Campesino, H., Team, T.M.T.S., Ciuffreda, L., Valenzuela-Fernández, A., and Flores, C. (2020). Fast SARS-CoV-2 detection by RT-qPCR in preheated nasopharyngeal swab samples. Int. J. Infect. Dis. https://doi.org/10.1016/j.ijid.2020.05.099

Broughton, J.P., Deng, X., Yu, G., Fasching, C.L., Servellita, V., Singh, J., Miao, X., Streithorst, J.A., Granados, A., and Sotomayor-Gonzalez, A. (2020). CRISPR–Cas12-based detection of SARS-CoV-2. Nat. Biotechnol., 1–5 https://doi.org/10.1038/s41587-020-0513-4

Brown, J.R., Atkinson, L., Shah, D., and Harris, K. (2020). Validation of an extraction-free RT-PCR protocol for detection of SARS-CoV2 RNA. medRxiv doi: https://doi.org/10.1101/2020.04.29.20085910

Bruce, E.A., Huang, M.-L., Perchetti, G.A., Tighe, S., Laaguiby, P., Hoffman, J.J., Gerrard, D.L., Nalla, A.K., Wei, Y., and Greninger, A.L. (2020). Direct RT-qPCR detection of SARS-CoV-2 RNA from patient nasopharyngeal swabs without an RNA extraction step. bioRxiv doi: 10.1101/2020.03.20.001008

Chen, J.S., Ma, E., Harrington, L.B., Da Costa, M., Tian, X., Palefsky, J.M., and Doudna, J.A. (2018). CRISPR-Cas12a target binding unleashes indiscriminate single-stranded DNase activity. Science 360, 436-439 10.1126/science.aar6245

Ding, X., Yin, K., Li, Z., Lalla, R.V., Ballesteros, E., Sfeir, M.M., and Liu, C. (2020). Ultrasensitive and visual detection of SARS-CoV-2 using all-in-one dual CRISPR-Cas12a assay. Nature communications 11, 1–10 https://doi.org/10.1038/s41467-020-18575-6

Döhla, M., Boesecke, C., Schulte, B., Diegmann, C., Sib, E., Richter, E., Eschbach-Bludau, M., Aldabbagh, S., Marx, B., and Eis-Hübinger, A.-M. (2020). Rapid point-of-care testing for SARS-CoV-2 in a community screening setting shows low sensitivity. Public Health https://doi.org/10.1016/j.puhe.2020.04.009

Fakheran, O., Dehghannejad, M., and Khademi, A. (2020). Saliva as a diagnostic specimen for detection of SARS-CoV-2 in suspected patients: a scoping review. Infectious diseases of poverty 9, 1–7 https://doi.org/10.1186/s40249-020-00728-w

Fozouni, P., Son, S., De León Derby, M.D., Knott, G.J., Gray, C.N., D’ambrosio, M.V., Zhao, C., Switz, N.A., Kumar, G.R., and Stephens, S.I. (2020). Direct detection of SARS-CoV-2 using CRISPR-Cas13a and a mobile phone. MedRxiv doi: https://doi.org/10.1101/2020.09.28.20201947

Gootenberg, J.S., Abudayyeh, O.O., Kellner, M.J., Joung, J., Collins, J.J., and Zhang, F. (2018). Multiplexed and portable nucleic acid detection platform with Cas13, Cas12a, and Csm6. Science 360, 439–444 DOI: 10.1126/science.aaq0179

Gootenberg, J.S., Abudayyeh, O.O., Lee, J.W., Essletzbichler, P., Dy, A.J., Joung, J., Verdine, V., Donghia, N., Daringer, N.M., and Freije, C.A. (2017). Nucleic acid detection with CRISPR-Cas13a/C2c2. Science 356, 438–442 DOI: 10.1126/science.aam9321

Grant, P.R., Turner, M.A., Shin, G.Y., Nastouli, E., and Levett, L.J. (2020). Extraction-free COVID-19 (SARS-CoV-2) diagnosis by RT-PCR to increase capacity for national testing programmes during a pandemic. BioRxiv doi: https://doi.org/10.1101/2020.04.06.028316

Hasan, M.R., Mirza, F., Al-Hail, H., Sundararaju, S., Xaba, T., Iqbal, M., Alhussain, H., Yassine, H.M., Perez-Lopez, A., and Tang, P. (2020a). Correction: Detection of SARS-CoV-2 RNA by direct RT-qPCR on nasopharyngeal specimens without extraction of viral RNA. PLoS One 15, e0240343

Hou, T., Zeng, W., Yang, M., Chen, W., Ren, L., Ai, J., Wu, J., Liao, Y., Gou, X., Li, Y., Wang, X., Su, H., Gu, B., Wang, J., and Xu, T. (2020). Development and evaluation of a rapid CRISPR-based diagnostic for COVID-19. PLoS Pathog. 16, e1008705. 10.1371/journal.ppat.1008705

Joung, J., Ladha, A., Saito, M., Segel, M., Bruneau, R., Huang, M.-L.W., Kim, N.-G., Yu, X., Li, J., and Walker, B.D. (2020). Point-of-care testing for COVID-19 using SHERLOCK diagnostics. N. Engl. J. Med., 1492–1494 DOI: 10.1056/NEJMc2026172

Jung, J., Garnett, E., Jariwala, P., Pham, H., Huang, R., Benzi, E., Issaq, N., Matzuk, M., Singh, I., and Devaraj, S. (2020). Clinical performance of a semi-quantitative assay for SARS-CoV2 IgG and SARS-CoV2 IgM antibodies. Clin. Chim. Acta 510, 790–795 https://doi.org/10.1016/j.cca.2020.09.023

Kellner, M.J., Koob, J.G., Gootenberg, J.S., Abudayyeh, O.O., and Zhang, F. (2019). SHERLOCK: nucleic acid detection with CRISPR nucleases. Nat. Protoc. 14, 2986–3012 https://doi.org/10.1038/s41596-019-0210-2

Kirby, T. (2021). New variant of SARS-CoV-2 in UK causes surge of COVID-19. The Lancet Respiratory Medicine DOI:https://doi.org/10.1016/S2213-2600(21)00005-9

Knott, G.J., and Doudna, J.A. (2018). CRISPR-Cas guides the future of genetic engineering. Science 361, 866–869 DOI: 10.1126/science.aat5011

Kriegova, E., Fillerova, R., and Kvapil, P. (2020). Direct-RT-qPCR Detection of SARS-CoV-2 without RNA Extraction as Part of a COVID-19 Testing Strategy: From Sample to Result in One Hour. Diagnostics 10, 605 https://doi.org/10.3390/diagnostics10080605

Lalli, M.A., Langmade, J.S., Chen, X., Fronick, C.C., Sawyer, C.S., Burcea, L.C., Wilkinson, M.N., Fulton, R.S., Heinz, M., Buchser, W.J., Head, R.D., Mitra, R.D., and Milbrandt, J. (2020). Rapid and Extraction-Free Detection of SARS-CoV-2 from Saliva by Colorimetric Reverse-Transcription Loop-Mediated Isothermal Amplification. Clin. Chem. 10.1093/clinchem/hvaa267

Li, Y., Li, S., Wang, J., and Liu, G. (2019). CRISPR/Cas systems towards next-generation biosensing. Trends Biotechnol. 37, 730–743 https://doi.org/10.1016/j.tibtech.2018.12.005

Merindol, N., Pépin, G., Marchand, C., Rheault, M., Peterson, C., Poirier, A., Houle, C., Germain, H., and Danylo, A. (2020). SARS-CoV-2 detection by direct rRT-PCR without RNA extraction. J. Clin. Virol. 128, 104423 doi: 10.1016/j.jcv.2020.104423

Meyerson, N.R., Yang, Q., Clark, S.K., Paige, C.L., Fattor, W.T., Gilchrist, A.R., Barbachano-Guerrero, A., and Sawyer, S.L. (2020). A community-deployable sars-cov-2 screening test using raw saliva with 45 minutes sample-to-results turnaround. medRxiv https://doi.org/10.1101/2020.07.16.20150250

Myhrvold, C., Freije, C.A., Gootenberg, J.S., Abudayyeh, O.O., Metsky, H.C., Durbin, A.F., Kellner, M.J., Tan, A.L., Paul, L.M., and Parham, L.A. (2018). Field-deployable viral diagnostics using CRISPR-Cas13. Science 360, 444–448 DOI: 10.1126/science.aas8836

Nguyen, L.T., Smith, B.M., and Jain, P.K. (2020). Enhancement of trans-cleavage activity of Cas12a with engineered crRNA enables amplified nucleic acid detection. Nature Communications 11, 4906 10.1038/s41467-020-18615-1

Ning, B., Yu, T., Zhang, S., Huang, Z., Tian, D., Lin, Z., Niu, A., Golden, N., Hensley, K., and Threeton, B. (2020). A smartphone-read ultrasensitive and quantitative saliva test for COVID-19. Science Advances 7, eabe3703 DOI: 10.1126/sciadv.abe3703

Ochert, A., Boulter, A., Birnbaum, W., Johnson, N., and Teo, C. (1994). Inhibitory effect of salivary fluids on PCR: potency and removal. PCR Methods Appl. 3, 365–368

Ostheim, P., Tichý, A., Sirak, I., Davidkova, M., Stastna, M.M., Kultova, G., Paunesku, T., Woloschak, G., Majewski, M., and Port, M. (2020). Overcoming challenges in human saliva gene expression measurements. Sci. Rep. 10, 1–12 https://doi.org/10.1038/s41598-020-67825-6

Patchsung, M., Jantarug, K., Pattama, A., Aphicho, K., Suraritdechachai, S., Meesawat, P., Sappakhaw, K., Leelahakorn, N., Ruenkam, T., and Wongsatit, T. (2020). Clinical validation of a Cas13-based assay for the detection of SARS-CoV-2 RNA. Nature Biomedical Engineering, 1–10 https://doi.org/10.1038/s41551-020-00603-x

Piepenburg, O., Williams, C.H., Stemple, D.L., and Armes, N.A. (2006). DNA detection using recombination proteins. PLoS Biol. 4, e204 https://doi.org/10.1371/journal.pbio.0040204

Procop, G.W., Shrestha, N.K., Vogel, S., Van Sickle, K., Harrington, S., Rhoads, D.D., Rubin, B.P., and Terpeluk, P. (2020). A direct comparison of enhanced saliva to nasopharyngeal swab for the detection of SARS-CoV-2 in symptomatic patients. J. Clin. Microbiol. 58 10.1128/JCM.01946-20

Ranoa, D., Holland, R., Alnaji, F.G., Green, K., Wang, L., Brooke, C., Burke, M., Fan, T., and Hergenrother, P.J. (2020). Saliva-based molecular testing for SARS-CoV-2 that bypasses RNA extraction. Biorxiv doi: https://doi.org/10.1101/2020.06.18.159434

Srivatsan, S., Han, P.D., Van Raay, K., Wolf, C.R., Mcculloch, D.J., Kim, A.E., Brandstetter, E., Martin, B., Gehring, J., and Chen, W. (2020). Preliminary support for a “dry swab, extraction free” protocol for SARS-CoV-2 testing via RT-qPCR. bioRxiv doi: 10.1101/2020.04.22.056283

Thai, H.T.C., Le, M.Q., Vuong, C.D., Parida, M., Minekawa, H., Notomi, T., Hasebe, F., and Morita, K. (2004). Development and evaluation of a novel loop-mediated isothermal amplification method for rapid detection of severe acute respiratory syndrome coronavirus. J. Clin. Microbiol. 42, 1956–1961 10.1128/JCM.42.5.1956-1961.2004

To, K.K.-W., Tsang, O.T.-Y., Yip, C.C.-Y., Chan, K.-H., Wu, T.-C., Chan, J.M.-C., Leung, W.-S., Chik, T.S.-H., Choi, C.Y.-C., and Kandamby, D.H. (2020). Consistent detection of 2019 novel coronavirus in saliva. Clin. Infect. Dis. https://doi.org/10.1093/cid/ciaa149

Uhm, J.-S., Ahn, J.Y., Hyun, J., Sohn, Y., Kim, J.H., Jeong, S.J., Ku, N.S., Choi, J.Y., Park, Y.-K., and Yi, H.-S. (2020). Patterns of viral clearance in the natural course of asymptomatic COVID-19: Comparison with symptomatic non-severe COVID-19. Int. J. Infect. Dis. 99, 279–285 https://doi.org/10.1016/j.ijid.2020.07.070

Vogels, C.B., Brito, A.F., Wyllie, A.L., Fauver, J.R., Ott, I.M., Kalinich, C.C., Petrone, M.E., Casanovas-Massana, A., Muenker, M.C., and Moore, A.J. (2020a). Analytical sensitivity and efficiency comparisons of SARS-CoV-2 RT–qPCR primer–probe sets. Nature microbiology 5, 1299–1305 https://doi.org/10.1038/s41564-020-0761-6

Vogels, C.B., Watkins, A.E., Harden, C.A., Brackney, D.E., Shafer, J., Wang, J., Caraballo, C., Kalinich, C.C., Ott, I.M., and Fauver, J.R. (2020b). SalivaDirect: A simplified and flexible platform to enhance SARS-CoV-2 testing capacity. Med https://doi.org/10.1016/j.medj.2020.12.010

Wee, S.K., Sivalingam, S.P., and Yap, E.P.H. (2020). Rapid direct nucleic acid amplification test without RNA extraction for SARS-CoV-2 using a portable PCR thermocycler. bioRxiv doi: 10.3390/genes11060664

Williams, E., Bond, K., Zhang, B., Putland, M., and Williamson, D.A. (2020). Saliva as a non-invasive specimen for detection of SARS-CoV-2. J. Clin. Microbiol. doi:10.1128/JCM.00776-20

Wyllie, A.L., Fournier, J., Casanovas-Massana, A., Campbell, M., Tokuyama, M., Vijayakumar, P., Warren, J.L., Geng, B., Muenker, M.C., and Moore, A.J. (2020). Saliva or nasopharyngeal swab specimens for detection of SARS-CoV-2. N. Engl. J. Med. 383, 1283–1286 DOI: 10.1056/NEJMc2016359

Xia, S., and Chen, X. (2020). Single-copy sensitive, field-deployable, and simultaneous dual-gene detection of SARS-CoV-2 RNA via modified RT–RPA. Cell Discovery 6, 1–4 https://doi.org/10.1038/s41421-020-0175-x

Zou, Y., Mason, M.G., and Botella, J.R. (2020). Evaluation and improvement of isothermal amplification methods for point-of-need plant disease diagnostics. PLoS One 15, e0235216 https://doi.org/10.1371/journal.pone.0235216

