## supplementary file for "A saliva-based RNA extraction-free workflow integrated with Cas13a for SARS-CoV-2 detection"

### Supplementary Figures

**Figure S1: Standardization of various primers for the detection of SARS-CoV-2**

**(A)** Detection of S gene amplicon by performing RT-qPCR using the SP-1 primers. The desired amplicon size was obtained under all the annealing temperatures tested (112bp). S gene plasmid was used as template. **(B)** Similarly, SP-4 primer for S gene amplification was used (134bp). **(C)** CDC-approved N-1 primer was used for PCR on N gene plasmid with a single amplicon (72bp) (d) PCR was performed using N-P1 primer which produced multiple amplicons. Black arrow head indicates the predicted amplicon size. The sequence of the respective primers is given in Supplementary Table 1.

**Figure S2: SHERLOCK validation of saliva samples using later-flow strips and signal quantitation method**

**(A1)** Paper-strip images of SARS-CoV-2 positive (*left*) and SARS-CoV-2 negative (*right*) samples which were subjected to SHERLOCK and LFA. Corresponding Ct values derived from RT-qPCR is shown above the Lateral-flow strip. The patient ID and Ct values are same as shown in *Figure 2f*. **(A2)** Corresponding T/C ratio of the images is shown. Red arrow head indicates the threshold T/C ratio **(B1)** Longitudinal test results of two patient saliva samples taken four times before the test results came negative. **(B2)** T/C ratio of the corresponding LFA images.

**Video 1:**

After opening the mobile application which is named as MI-SEHAT, the user will be directed to enter the details like age and name (optional). For COVID-19 test, user will click on the tab "COVID-19 test" which will open the mobile phone camera. For simplicity we have used a dummy cassette which will hold the test strip and trained the software along with the cassette. The user will select a region which should include the blue mark, and the two lines below the area where CASSPIT is written. After clicking the image, the software will display the result, which in the given example are positive, as only two bands are detected one in the control lane and another in the test lane. The user will have an option to save the test results for future use or to consult the team of experts regarding any query related to the assay. The software has also provision to provide a signal quantitation of the test results which are displayed as score. In positive samples the test score above 0.15.

**Video 2:**

This is an example of a negative test result, where only one band can be seen (control lane). The test score is also displayed, which in case of negative samples will be below 0.15.

**Table S1: List of primers, probes, crRNA, and synthetic DNA fragments**

| Name | Sequence | Sources | Note | REF |
|---|---|---|---|---|
| **RPA-and IVT** | | | | |
| T7 S-gene Fw | 5’**gaaattaatacgactcactatagggAGGTTTCAA ACTTTACTTGCTT TACATAGA-3’** | ILS | Lower-case letters indicate overhang T7 promotor sequence | Zhang Lab protocols, MIT |
| T7 S-gene Rev | 5’-TCCTAGGTTGAAGATAACCCACATAAT AAG-3’ | ILS | | Zhang Lab protocols, MIT |
| T7-Orf1ab Fw | 5’**gaaattaatacgactcactataggg**CGAAGTTGT AGGAGACATTAT ACTTAAACC-3’ | ILS | Lower-case letters indicate overhang T7 promotor sequence | Zhang Lab protocols, MIT |
| T7-Orf1ab Rev | 5’-TAGTAAGACTAGAATTGTCTACATAAG CAGC-3’ | ILS | | Zhang Lab protocols, MIT |
| T7-S-FP | **5’-gaaattaatacgactcactataggg**TTTTTCGGCTT -3’ | Sigma | Lower-case letters indicate overhang T7 promotor sequence | This study |
| T7-S-RP | 5’TCTAACAATAGATTCTGTTGGTTGGA CTCTAAAGTT-3’ | | | This study |
| T7-Orf1ab FP | 5’-gaaattaatacgactcactatagggAATGGATAAT-3’ | Sigma | | This study |
| T7-Orf1a RP | 5’-TTAGCTATAGTATCCCAAGGGACACTA TTAACA-3’ | | | This study |
| **crRNA** | | | | |
| S gene | 5’**gauuuagacuaccccaaaaacgaaggggacuaaaa c**GCAGCACCAGCUGUCCAACCUGAAG AAG-3’ | Synthego | Lower-case letters indicate scaffold sequence | Zhang Lab protocols, MIT |
| Orf1ab | 5’**gauuuagacuaccccaaaaacgaaggggacuaaaa cc**CAACCUCUUCUGUAAUUUUUAAACU | Synthego | Lower-case letters | Zhang Lab protocols, |

| | | | | |
|---|---|---|---|---|
| | AU-3’ | | indicate scaffold sequence | MIT |
| **Reporter** | | | | |
| Cas13a Reporter | 5’/56FAM/mArArUrGrGrCmAmArArUrGrG rCmA/3Bio/-3’ | IDT | Lateral Flow Reporter | |
| **Primer sequences** | | | | |
| SP-1 (FP) | AGGTTTCAAACTTTACTTGCTTTA CATAGA | | Used for PCR | This study |
| SP-1 (RP) | CTTATTATGTGGGTTATCTTCAAC CTAGGA | | Used for PCR | |
| SP-2 (FP) | ACTGTGCACTTGACCCTCTCT | Bio serve | Used for PCR | This study |
| SP-2 (RP) | CACCAAAAGGGCACAAGTTTGTA A | Bio serve | Used for PCR | This study |
| SP-3 (FP) | TGACCCTCTCTCAGAAACAAAGT GT | Bio serve | Used for PCR | This study |
| SP-3 (RP) | AACTTCACCAAAAGGGCACAAGT | Bio serve | Used for PCR | This study |
| SP-4 (FP) | 5’- ACTGTGCACTTGACCCTCTCTCAG -3’ | ILS | Used for PCR | This study |
| SP-4 (FP) | 5’- AGTTTGTAATATTAGGAAATCTA- 3’ | ILS | Used for PCR | This study |
| NP-1 (FP) | 5’-ACCCGCAATCCTGCTAACAA-3’ | Bio serve | Used for PCR | This study |
| NP-1 (RP) | 5’-ACGAGAAGAGGCTTGACTGC-3’ | Bio serve | Used for PCR | This study |
| NP-2 (FP) | 5’-ATCACATTGGCACCCGCAAT-3’ | Bio serve | Used for PCR | This study |
| NP-2 (RP) | 5’-GTTGCGACTACGTGATGAGGA- 3’ | Bio serve | Used for PCR | This study |
| NP-3 (FP) | 5-’CACATTGGCACCCGCAATC-3’ | ILS | Used for PCR | This study |
| NP3 (RP) | 5’-GAGGAACGAGAAGAGGCTTG- 3’ | ILS | Used for PCR | This study |
| N1 (FP) | 5’-GACCCCAAAATCAGCGAAAT-3’ | Sigma | Used for PCR | Chantal et al., 2020 |
| N1 (RP) | 5’- TCTGGTTACTGCCAGTTGAATCTG -3’ | Sigma | Used for PCR | Vogels et al., 2020 |
| Orf1ab-FP | 5’- AGGAGACATTATACTTAAACCAG | Sigma | Used for PCR | This study |

| | | | | |
|---|---|---|---|---|
| | CA-3’ | | | |
| Orf1ab-RP | 5’-TAGATCTGTGTGGCCAACCTC-3’ | Sigma | Used for PCR | This study |
| Orf1ab-V-FP | 5’-CCCTGTGGGTTTTACACTTAA-3’ | Sigma | Used for PCR | This study |
| Orf1ab-V-RP | 5’-ACGATTGTGCATCAGCTGA-3’ | Sigma | Used for PCR | This study |
| RNase FP | 5’-AGATTTGGACCTGCGAGCG-3’ | Sigma | Used for PCR | |
| RNase RP | 5’-GAGCGGCTGTCTCCACAAGT-3’ | Sigma | Used for PCR | |
| **PROBES** | | | | |
| SP-1 Probe sequence | FAM-CTCCTGGTGATTCTTCTTCAGG BBQ | Sigma | Used for PCR | |
| SP-1, SP-2, SP-3 Probe sequence | 6FAM-AATCTATCAAACTTCTAACTTTA-BBQ | ILS | Used for PCR | This study |
| NP-1, NP-2, and NP3 Probe Sequence | 6FAM-ACTTCCTCAAGGAACAACATTGCCA-BBQ | ILS | Used for PCR | This study |
| N1 Probe sequence | FAM-ACCCCGCATTACGTTTGGTGGACC - BBQ | Sigma | Used for PCR | Vogels et al., 2020 |
| RNAse P | FAM-TTCTGACCTGAAGGCTCTGCGCG-BBQ | Sigma | Used for PCR | Vogels et al., 2020 |

| | | | | |
|---|---|---|---|---|
| **Template sequence for IVT** | | | | |
| S-synthetic fragment 1 | **gaaattaatacgactcactatagggg**AGGTTTCAAACTTTACTTGCTTTACATAGAAGTTATTTGACTCCTGGTGATTCTTCTTCAGGTTGGACAGCTGGTGCTGCAGCTTATTATGTGGGTTATCTTCAACCTAGGACTT | Xcelris | Used for IVT | This study |
| S synthetic fragment 2 | **gaaattaatacgactcactatagggg**ACTGTGCACTTGACCCTCTCTCAGAAACAAAGTGTACGTTGAAATCCTTCACTGTAGAAAAAGGAATCTATCAAACTTCTAACTTTAGAGTCCAACCAACAGAATCTATTGTTAGATTTCCTAATATTACAAACTTGTGCCCTTTTGGTGAAGTT | Xcelris | Used for IVT | This study |
| Orf1ab | **gaaattaatacgactcactatagggg**CTACCGA | Xcelris | Used for | This |

| | | | | |
|---|---|---|---|---|
| synthetic fragment | AGTTGTAGGAGACATTATACTTA AACCAGCAAATAATAGTTTAAAA ATTACAGAAGAGGTTGGCCACAC AGATCTAATGGCTGCTTATGTAG ACAATTCTAGTCTTACTATTAA | | IVT | study |

Abbreviations: FP: Forward Primer; RP: Reverse primer; Fw: Forward sequence; Rev: Reverse sequence. SP: S gene primer; NP: N gene primer; N1: USA CDC approved primer for N gene; RPA: Recombinase polymerase amplification; IVT: Invitro transcription.

**References**

Vogels, C. B. F., Brito, A. F., Wyllie, A. L., Fauver, J. R., Ott, I. M., Kalinich, C. C., et al. (2020a). Analytical sensitivity and efficiency comparisons of SARS-CoV-2 RT–qPCR primer–probe sets. Nat. Microbiol. doi:10.1038/s41564-020-0761-6.

Link to the Zang Lab protocol

https://www.broadinstitute.org/files/publications/special/COVID-19%20detection%20(updated).pdf
